# Impact of Life’s Essential 8 Cardiovascular Health Score and Hypertension on Retinal Biological Aging

**DOI:** 10.1101/2025.10.16.25338198

**Authors:** David Squirrell, Christopher Nielsen, Ehsan Vaghefi, Songyang An, Shima Moghadam, Li Xie, Michael V. McConnell

## Abstract

**Background:** The American Heart Association’s Life’s Essential 8 (LE8) score was designed to quantify an individual’s cardiovascular health (CVH). Previously we developed a novel biomarker (retinal BioAge) to estimate biological aging from a deep-learning analysis of retinal images. This study investigates the association between retinal BioAge and CVH, as quantified by LE8, and identifies the key health factors driving this relationship.

**Methods:** This retrospective study included 4,887 participants (aged 40-70 years) from the UK Biobank. The retinal BioAgeGap (retinal BioAge minus chronological age) was calculated from retinal images. A CVH score was computed for each participant.

**Results:** Participants in the highest CVH quartile were shown to have a negative retinal BioAgeGap while the lowest CVH quartile had a positive retinal BioAgeGap (-0.36 vs. +0.30 years; difference = - 0.65 years, P<0.001). Ablation analysis identified blood pressure as the most significant driver of this association, accounting for 66.3% of the total change in effect size (∣Δd∣=0.11, P<0.001). Among individuals with high CVH scores, a negative retinal BioAge was no longer found among those with hypertension (≥130/80mmHg) compared to those without (+0.08 vs -0.52 years, P<0.001).

**Conclusions:** Better CVH is strongly associated with a “younger” retinal BioAge. Blood pressure was the predominant contributor to this association and an accelerated retinal BioAge was noted even in those individuals who had an otherwise favorable CVH score but were hypertensive. This study highlights retinal BioAge’s potential as a scalable, non-invasive tool for refining cardiovascular risk assessment.

## Introduction

Cardiovascular disease (CVD) remains the leading cause of mortality worldwide, creating a significant public health and economic burden. [1, 2] In response, a global emphasis has been placed on preventive health strategies to mitigate CVD risk factors before the onset of clinical disease. [3] Central to this effort is the American Heart Association’s (AHA) Life’s Essential 8 (LE8) score, a comprehensive metric designed to promote and quantify an individual’s cardiovascular health (CVH). [4] The LE8 score is composed of eight key health factors and behaviors: Diet, Physical activity, Nicotine exposure, Sleep health, Body mass index (BMI), Blood lipids (non-HDL cholesterol), Blood glucose (HbA1c), and Blood pressure (BP). A favorable LE8 score is strongly associated with a lower risk of incident CVD, stroke, heart failure, and all-cause mortality, establishing it as a powerful tool for public health and clinical practice. [5, 6]

The human retina offers a unique and non-invasive window into the body’s microvasculature. Retinal imaging, a routine component of eye care, can reveal pathological changes that reflect systemic vascular health long before symptoms manifest. [7, 8] “End organ” changes within the retinal microvasculature have been linked to hypertension, diabetes, and an increased risk of stroke and coronary heart disease. [9, 10] Recent advancements in artificial intelligence (AI), particularly deep learning (DL), have enabled the development of models that can analyze retinal images to predict a wide range of systemic health indicators. [11-14] Building on this, our group recently developed and validated a DL model to predict 10-year atherosclerotic CVD (ASCVD) risk directly from retinal images. [14] This work has been extended to derive a novel biomarker: retinal BioAge, which we have shown correlates with leukocyte telomere length and cardiovascular risk factors. [15]

This study aims to investigate the relationship between the non-invasively derived retinal BioAge and an individual’s CVH, as quantified by their LE8 score. By analyzing this association in a large dataset, we seek to understand if CVH behaviors and factors have a measurable impact on retinal biological aging. It also seeks to identify which individual LE8 factors, such as blood pressure, are the most significant contributors to this association. Importantly, retinal imaging has advantages as a promising, scalable tool for identifying at-risk individuals, thereby enabling earlier and more targeted lifestyle and pharmacological interventions to slow biological aging and prevent cardiovascular disease.

## Methods

### Dataset

This retrospective, cross-sectional study utilized data from the UK Biobank (UKBB), a large-scale biomedical database containing in-depth health information from half a million UK participants. [16] As described in our prior work, [15] we analyzed a cohort of 51,955 participants with a pair of acceptable-quality macula-centered retinal images. In total, data from 43,349 UKBB participants were used to train the retinal BioAge model and a separate cohort of 5,476 UKBB participants was used for testing. For this current work, we further refined the test set of 5,476 participants to exclude those who did not have a BMI recorded. The final analytical cohort therefore comprised 9,774 retinal images from 4,887 participants aged 41-70 years (Figure 1). The demographic and relevant biometric data from these participants together with the original test set are presented in Table 1.

**Table 1:** The demographic and risk factor composition of the 5476 individuals that formed the original test dataset of the retinal BioAge deep-learning model [15] and the subset of 4887 individuals analyzed for the test dataset in this study. Significance testing was performed between the original retinal BioAge test dataset and the test dataset used in this study after excluding patients with missing values (see Figure 1). There was no significant difference across all demographics and risk factors.

|  |  | Original retinal BioAge test dataset<br>(N= 5,476) |  | Test dataset analyzed in this study –<br>a subset of the original retinal<br>BioAge study<br>(N= 4,887) |  |
| --- | --- | --- | --- | --- | --- |
|  |  | Mean | Std | Mean | Std |
| Age (years) |  | 56.1 | 8.0 | 56.0 | 8.0 |
| Systolic Blood Pressure<br>(mmHg) |  | 135.3 | 17.2 | 135.2 | 17.2 |
| Diastolic Blood Pressure<br>(mmHg) |  | 81.2 | 9.6 | 81.2 | 9.7 |
| Hemoglobin A1c (%) |  | 5.4% | 0.5% | 5.4% | 0.5% |
| Total cholesterol(mg/dL) |  | 219.9 | 42.8 | 220.1 | 42.9 |
| HDL cholesterol(mg/dL) |  | 162.2 | 40.2 | 57.8 | 15.1 |
| BMI |  | 27.0 | 4.6 | 27.0 | 4.6 |
| Urine Albumin to<br>Creatinine Ratio (mg/g) |  | 29.8 | 131.2 | 31.0 | 139.1 |
| estimated Glomerular<br>Filtration Rate<br>(mL/min/1.73 m<sup>2</sup>) |  | 96.1 | 14.1 | 96.1 | 14.0 |
| Sex assigned at birth |  | Male | Female | Male | Female |
|  |  | 2,470 (45%) | 3,006 (55%) | 2,206<br>(45%) | 2,681<br>(55%) |
| Current Smoker |  | TRUE | FALSE | TRUE | FALSE |
|  |  | 459<br>(8.4%) | 4,999 (91.6%) | 428<br>(8.8%) | 4,459<br>(91.2%) |
| Diabetes (%) |  | 247<br>(5%) | 5,213<br>(95%) | 216<br>(5%) | 4,660<br>(95%) |
| Race and<br>Ethnicity | Black | 2% |  | 2% |  |
|  | East Asian | <1% |  | <1% |  |
|  | Non-Hispanic<br>White | 94% |  | 94% |  |
|  | South Asian | 2% |  | 2% |  |
|  | Multi-racial | <1% |  | <1% |  |
|  | Other | 1% |  | 1% |  |
|  | Declined/<br>Unknown | 1% |  | 1% |  |

**Figure 1:**
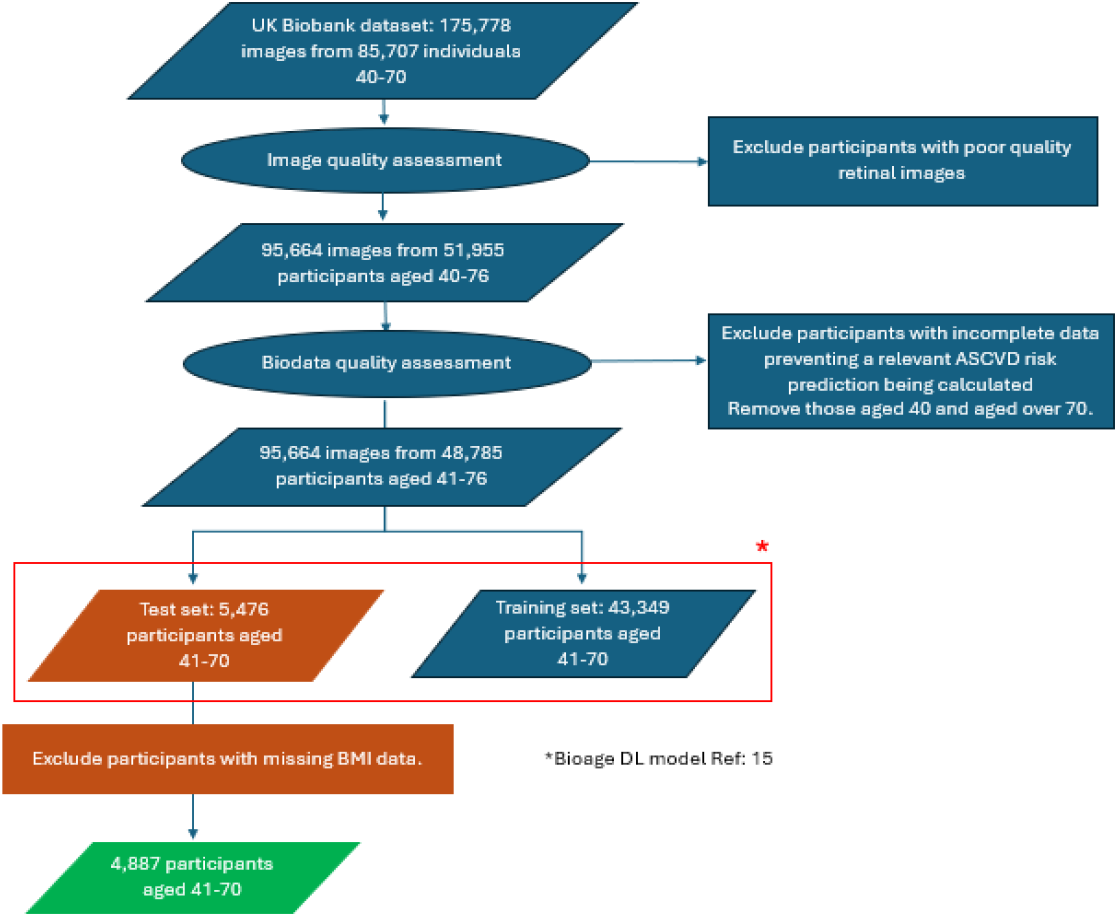
Flow chart illustrating the composition of the dataset used in this study.

### Ethics statement

Our research adhered to the protocols and approvals governed by UKBB. As published previously, our analysis of this de-identified data set was approved by University of Auckland (UoA 86299), to ensure the responsible use and secure transfer of data.

### Retinal BioAge and BioAgeGap Calculation

The retinal BioAge DL model was developed as an extension of a previously published model that predicts 10-year ASCVD risk from retinal fundus images. [14] The methodology has been detailed elsewhere. [15] In brief, the process involves several steps:

1. DL model extracts multi-dimensional features from an individual’s retinal image to predict their 10-year ASCVD risk score.
2. This AI-predicted ASCVD risk score is compared to the distribution of ASCVD risk scores from other individuals of varying chronological ages in a large training cohort.
3. The model identifies a group of “closest neighbors” whose retinal features and associated ASCVD risk profiles are most similar to the individual’s.
4. The retinal BioAge is then calculated as the mean chronological age of these closest neighbors, weighted probabilistically.
5. Finally, the retinal BioAgeGap is computed for each participant by subtracting their chronological age from their calculated retinal BioAge: BioAgeGap = Retinal BioAge − Chronological Age

To control for the inherent correlation between chronological age and the retinal BioAgeGap, we implemented an age-bin normalization procedure. Participants were grouped into 5-year age bins (41–45, 46–50, etc.). The mean BioAgeGap was calculated for each bin, and this mean was then subtracted from each individual participant’s raw BioAgeGap. This produced a normalized BioAgeGap, which represents the deviation from the average for an individual’s age group, effectively isolating individual biological aging differences.

### Cardiovascular Health (CVH) Score Calculation

As three of the components of the AHA LE8 scores (Diet, Physical activity, and Sleep) were not available in our UKBB dataset, the CVH score used in this study was based on the five available LE8 components: Nicotine exposure, BMI, Non-HDL cholesterol, HbA1c, BP. Per AHA’s CVH scoring methodology, [4] scores of 0 to 100 for each of the five LE8 components were determined and the CVH score was calculated as the sum of these five component scores divided by five (with the resulting range also 0 to 100).

### Statistical Analysis

To investigate the relationship between CVH health and retinal BioAge, we first stratified participants into quartiles based on their CVH score. We then compared the mean normalized retinal BioAgeGap between the least healthy participants (i.e., bottom CVH score quartile, Q1) and most healthy participants (top CVHscore quartile, Q4). To account for unequal variances, this comparison was performed using Welch’s t-test.

To determine the relative contribution of individual LE8 factors to the observed association, we conducted an ablation study by systematically removing each of the five components (BMI, Non-HDL cholesterol, HbA1c, BP). Having removed the component under test, the CVH score was recalculated and the corresponding Cohen’s d effect size for the top-versus-bottom quartile comparison. The importance of each component was quantified by the change in effect size (Δd) after its removal. A negative Δd would indicate that removing the component under test weakened the association between the CVH score and BioAgeGap. To determine statistical significance for this change, we employed a non-parametric bootstrap procedure with 1,000 resamples to generate 95% CIs and a two-tailed p-value for each component’s Δd.

All statistical analyses were performed using Python (version 3.9) with the pandas, numpy, and SciPy libraries, and statistical significance was set at p < 0.05.

## Results

### Association Between mLE8 Score and Retinal BioAgeGap

There was a strong and significant association between an individual’s CVH score and their retinal BioAgeGap for all participants across age bins (Figure 2). Participants in the bottom CVH score quartile exhibited accelerated retinal aging, with a mean normalized retinal BioAgeGap of +0.30 (95CI 0.16-0.44) years. In contrast, those in the top CVH score quartile showed decelerated ageing, with a mean retinal BioAgeGap of -0.36 (95%CI -0.52, -0.20) years. The difference of 0.65 years between the healthiest and least healthy quartiles was significant (*P*<0.001). This trend was observed in both males and females, although the effect was more pronounced in males (difference of 0.87 years vs. 0.56 years for females) (Table 2).

**Table 2.** Comparison of Retinal BioAgeGap Between Lowest and Highest Cardiovascular Health (CVH) Score Quartiles.

| Subgroup | BioAgeGap for<br>Lowest CVH Score<br>Quartile<br>(Mean, 95% CI) | BioAgeGap for<br>Highest CVH<br>Score Quartile<br>(Mean, 95% CI) | BioAgeGap<br>Difference<br>(Mean, 95% CI) | P-value |
| --- | --- | --- | --- | --- |
| All<br>Participants | 0.30 (0.16, 0.44) | -0.36 (-0.52, -0.20) | 0.65 (0.86, 0.44) | <0.001 |
| Male | 0.40 (0.19, 0.61) | -0.47 (-0.71, -0.23) | 0.87 (1.12, 0.55) | <0.001 |
| Female | 0.28 (0.08, 0.487) | -0.29 (-0.51, -0.07) | 0.56 (0.86, 0.27) | <0.001 |

**Figure 2.**
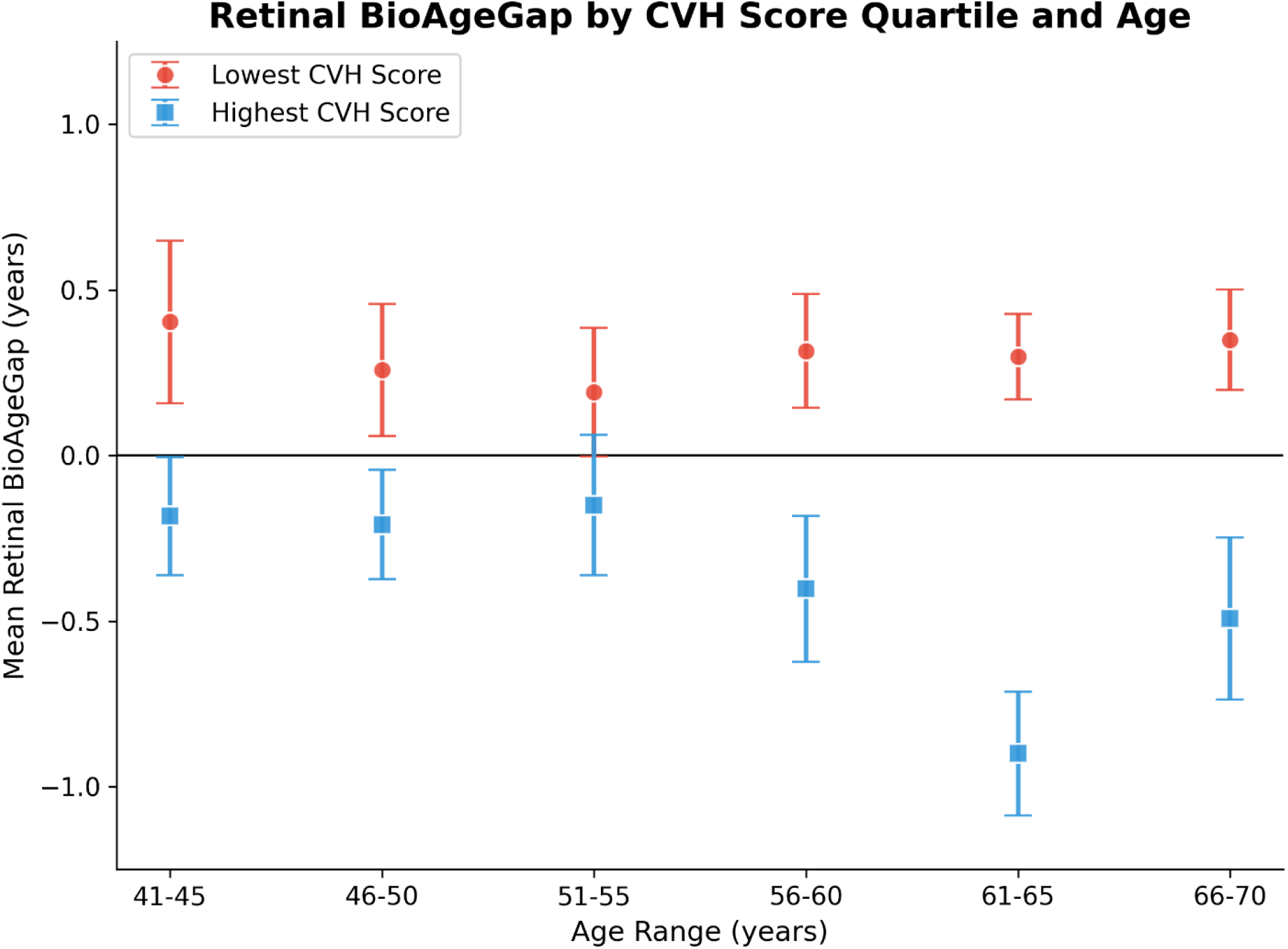
Retinal BioAgeGap by Cardiovascular Health (CVH) Score Quartile and Age. The figure shows retinal BioAgeGap means and 95% confidence intervals across age bins (41-45 to 66-70 years) for the lowest CVH score quartile (red) and the highest CVH score quartile (blue), demonstrating consistent separation across age groups.

### Relative Importance of LE8 Components

Ablation analysis revealed that BP was the most significant LE8 contributor to the association between the CVH score and retinal BioAgeGap (Figure 3). Removing BP from the CVH score resulted in the largest reduction in the observed effect size. The negative absolute change in Cohen’s d (∣Δd∣) -0.11 (95% CI: 0.07 to 0.16; p<0.001) indicates that the exclusion of BP substantially weakened the observed relationship between CVH status and accelerated retinal aging. Blood cholesterol had the next largest impact, but this trend did not reach statistical significance (∣Δd∣=0.04, p=0.12). The contributions of BMI (∣Δd∣=0.01, p=0.82), blood glucose (∣Δd∣<0.01, p=0.74), and nicotine exposure (∣Δd∣<0.01, p=0.68) were minimal (Table 3).

**Table 3.** Contribution of the individual components of the composite mLE8 to the relationship between CVH score and retinal BioAgeGap as revealed by variable importance analysis. BP accounts for 66.3% of the effect and is the dominant influence on this relationship.

| Variable | Absolute Effect Size Delta<br>\|Δd\| (years) | % of Total Change in<br>Effect Size |  |
| --- | --- | --- | --- |
| Blood Pressure | 0.11 | 66.3% | p<0.001 |
| Blood Lipids | 0.04 | 21.3% | P=0.12 |
| BMI | 0.01 | 5.6% | P=0.82 |
| Blood Glucose | <0.01 | 3.6% | P=0.78 |
| Nicotine Exposure | <0.01 | 3.2% | P=0.68 |

**Figure 3.**
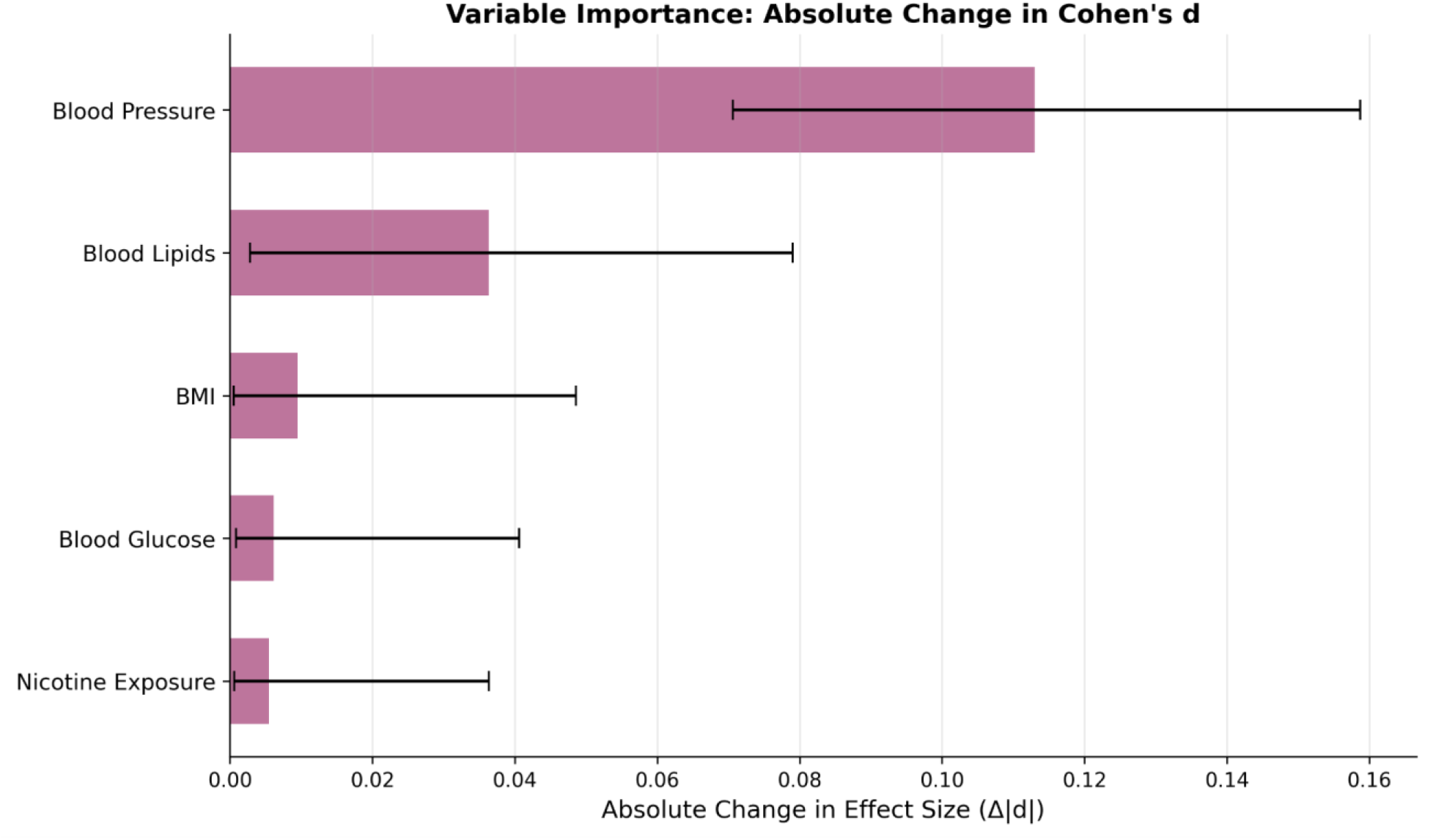
Variable importance analysis to evaluate the influence of the individual components of the mLE8 CVH score to an individual’s retinal BioAgeGap. This revealed that removing BP led to the largest absolute change in effect size (Δ|d|) for the impact of CVH score on retinal BioAgeGap. Error bars show 95% confidence intervals. The percentage contribution of each variable to the overall change in effect size can be seen in Table 3. BMI – Body Mass Index.

Having identified that BP had the most important contribution to the association between CVH score and retinal BioAgeGap, a further analysis was conducted to assess the specific impact of hypertension on this association. Participants in the high CVH score and low CVH score quartiles were therefore subdivided based on whether their BP was under or above the clinical threshold of ≥130/80 mm Hg set out in the latest AHA hypertension guidelines [17]. This created four distinct analytical groups: (1) High CVH / not hypertensive, (2) High CVH / hypertensive, (3) Low CVH / not hypertensive, and (4) Low CVH / hypertensive. We then calculated the mean retinal BioAgeGap and 95% confidence interval (CI) for each group. We found that the group with more optimal CVH, i.e., High CVH /not hypertensive had a significantly better retinal BioAgeGap (-0.52 years [-0.72 to -0.33]) than the group with the poorest CVH profile, i.e., Low CVH / hypertensive (+0.44 years [+0.29 to +0.59]; p<0.001). The intermediate groups demonstrated a graded, but equally significant relationship (Figure 4). Finally, to isolate the specific effect of hypertension among individuals with otherwise good CVH, an independent samples t-test was used to compare the retinal BioAgeGap between the High CVH / not hypertensive and High CVH / hypertensive groups. Although all individuals in this analysis had good CVH scores, the retinal BioAgeGap in those individuals with hypertension was significantly worse than those without hypertension (-0.52 years [-0.72 to -0.33] vs. +0.08 years [-0.20 to +0.35]; p<0.001).

**Figure 4.**
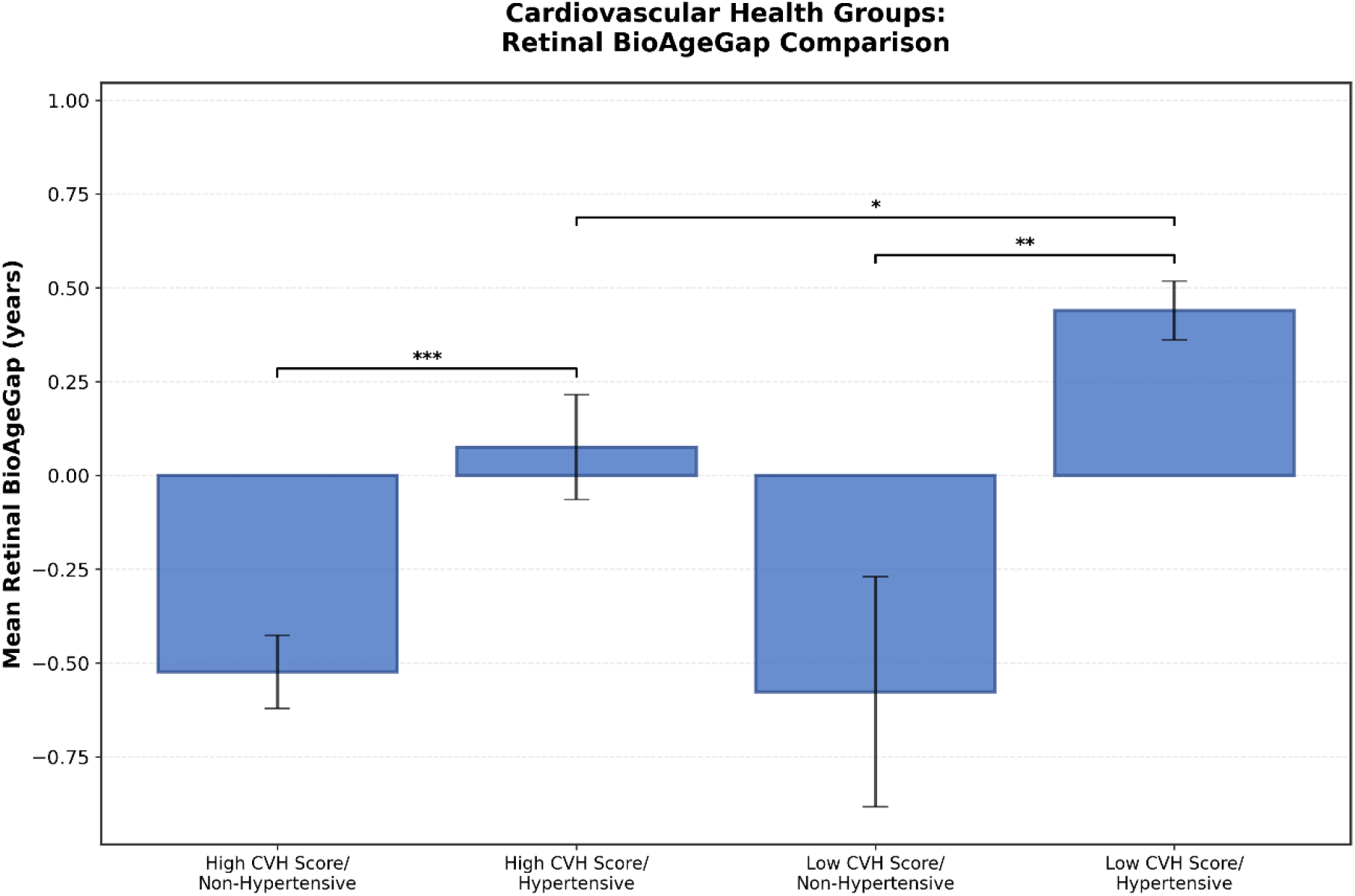
Bar chart showing comparison of retinal BioAgeGap in different CVH groupings by CVH score and hypertension status. The asterisks (*) above the bars indicate statistical significance levels: *** = p < 0.001, ** = p < 0.01, and * = p < 0.05.

## Discussion

This study demonstrates a significant association between an individual’s CVH, as quantified by the AHA’s LE8 score [3] and an individual’s retinal BioAge, derived non-invasively from retinal images. Individuals with better CVH scores exhibited a significantly lower retinal BioAgeGap compared to those with worse CVH scores. The retinal vascular network shares embryological origins, anatomical features, and physiological properties with the cerebral and coronary microcirculations, making it an accessible surrogate for assessing systemic vascular health. [18,19, 20] Our results align with this body of evidence and indicate that changes in the retina could be a useful proxy for CVH, and hypertension in particular. The 0.65-year difference in the retinal BioAgeGap between the highest and lowest CVH quartiles observed in this study provides a quantifiable measure of this association.

The ablation analysis revealed that the dominant contributions to the observed association between retinal BioAgeGap and CVH was BP, with Non-HDL cholesterol playing a less dominant role. Removing BP from the CVH score calculation caused the effect size to diminish by an order of magnitude more than any other component, highlighting the profound sensitivity of the retina to elevated BP. Chronic elevation in BP inflicts direct mechanical stress on vessel walls, leading to endothelial dysfunction, remodeling, arteriolar narrowing, and increased vessel stiffness, [21,22] features that our DL model likely interprets as signs of accelerated cardiovascular aging. This aligns with foundational studies like the Atherosclerosis Risk in Communities (ARIC) study, which have shown strong links between elevated blood pressure and adverse retinal vascular measurements [23] and has significant implications for cardiovascular risk stratification and future preventative health strategies.

Subgroup analysis revealed that not only was BP the primary driver of the relationship between the CVH score and retinal BioAgeGap, but “younger” retinal BioAge seen with good CVH scores was negated in participants whose BP was above the AHA hypertension threshold, even when overall CVH scores were similar. [17] These findings suggest that although the LE8 score is a powerful public health tool, it may not fully capture risk in “healthy but hypertensive” individuals, so retinal BioAge could be a useful adjunct providing a more biological indicator of vascular health that is sensitive to changes in the retina resulting from hypertension.[24]. Indeed, both US and European hypertension guidelines highlight the retina for assessment of target organ damage to guide therapy.[17, 24]

The findings of this study have significant potential for clinical translation. We have previously shown that our retinal BioAge DL algorithm can identify individuals at higher risk for key indicators of cardiovascular-kidney-metabolic (CKM) syndrome compared to their chronological peers. [25] We have also demonstrated that an elevated retinal BioAge is inversely associated with leukocyte telomere length, suggesting that retinal images may provide a proxy for these fundamental aging processes at the cellular level. [15] The ability to calculate a retinal BioAge from a routine fundus photograph, a rapid, non-invasive, and increasingly automated procedure, therefore offers a scalable method for opportunistic screening in both ophthalmic and primary care settings. [26, 27] Our results suggest that retinal BioAge could serve as a novel community screening tool identifying those individuals whose overall health profile appears favorable but who may benefit from more aggressive blood pressure surveillance and management. An accelerated retinal BioAge could also serve as a powerful, personalized “call to action,” raising awareness of the importance of potential elevated CKM risk, and hypertension in particular, and help initiate conversations about lifestyle modifications or pharmacological interventions. Finally, this tool could also help motivate behavioral change by making the abstract concept of “risk” more tangible through the intuitive metric of “retinal BioAge.” [28]

This study has several limitations. First, its cross-sectional design prevents us from inferring causality; we can only report associations between CVH scores and the retinal BioAgeGap at a single point in time. Second, the CVH score was derived from the five (out of the eight) LE8 lifestyle and biomarker components available to us. While this excludes lifestyle factors like diet and physical activity, it ensures the observed association is driven by objective physiological data. Incorporating the full suite of LE8 metrics in future research represents an opportunity to build upon and potentially strengthen the current findings. Third, the study cohort was drawn from UKBB, which is predominantly composed of individuals of European ancestry. [16] This limits the generalizability of our findings to more diverse racial and ethnic populations, which often bear a disproportionate burden of cardiovascular disease. [29]

Future research should focus on validating these findings in longitudinal diverse cohorts to establish a causal relationship between changes in LE8 components (particularly BP) and the trajectory of retinal BioAgeGap over time. Prospective studies are also needed to determine if interventions that lower retinal BioAgeGap lead to a reduction in incident cardiovascular events.

## Conclusion

In summary, this study establishes a strong, graded association between an individual’s cardiovascular health and their retinal BioAge in a large cohort from the UKBB. Compared to their peers, individuals with better cardiovascular health, as assessed by the AHA’s Life’s Essential 8 score, had a “younger” retinal BioAge, while those with poorer cardiovascular health were found to have an “older” retinal BioAge. We also demonstrate that blood pressure was the dominant factor influencing this relationship. Our findings highlight a crucial clinical gap: namely individuals may have an otherwise healthy cardiovascular profile but still harbor inadequately controlled blood pressure. The retinal BioAge emerges as a highly sensitive and accessible biomarker capable of detecting this risk, offering a powerful, non-invasive tool to refine cardiovascular risk assessment and motivate timely preventive interventions. This approach could significantly enhance public health strategies aimed at mitigating the global burden of cardiovascular disease.

## Data Availability

All data used in this manuscript is publicly available through the UK Biobank.

## References

[1] Roth, G. A., Mensah, G. A., Johnson, C. O., et al. (2020). Global Burden of Cardiovascular Diseases and Risk Factors, 1990–2019: Update From the GBD 2019 Study. Journal of the American College of Cardiology, 76(25), 2982–3021.

[2] World Health Organization. (2021). Cardiovascular diseases (CVDs). https://www.who.int/news-room/fact-sheets/detail/cardiovascular-diseases-(cvds)

[3] Nabaty, N.L., Menon, T., Trang, G., Vijay, A., Chogyal, L., Cataldo, R., Govind, N., Jain, P., Singh, P., Dolasa, N., Sahani, M., Deedwania, P. and Vijayaraghavan, K. (2024) ‘Global Community Health Screening and Educational Intervention for Early Detection of Cardiometabolic Renal Disease’, Annals of Global Health, 90(1), p. 54. Available at: 10.5334/aogh.4497

[4] Hong, S., Wang, Y., Song, Y., et al. (2023). Life’s Essential 8 and the risk of all-cause and cause-specific mortality: a prospective study of 486,337 adults. BMC Medicine, 21(1), 195.

[5] GBD 2019 Risk Factors Collaborators. (2020). Global burden of 87 risk factors in 204 countries and territories, 1990–2019: a systematic analysis for the Global Burden of Disease Study 2019. The Lancet, 396(10258), 1223–1249.

[6] Kelly, B. B., Narula, J., & Fuster, V. (2012). Promoting global cardiovascular health. Nature Reviews Cardiology, 9(12), 738–741.

[7] London, A., Benhar, I., & Schwartz, M. (2013). The retina as a window to the brain—from eye research to CNS disorders. Nature Reviews Neurology, 9(1), 44–53.

[8] Wagner, S.K., Fu, D.J., Faes, L., Liu, X., Huemer, J., Khalid, H., Ferraz, D., Korot, E., Kelly, C., Balaskas, K. and Denniston, A.K., 2020. Insights into systemic disease through retinal imaging-based oculomics. Translational vision science & technology, 9(2), pp.6–6.

[9] Wong, T. Y., Klein, R., Klein, B. E. K., et al. (2001). Retinal microvascular abnormalities and risk of mortality from cardiovascular disease: the Beaver Dam Eye Study. Ophthalmology, 108(9), 1630–1636.

[10] Mitchell, P., Wang, J. J., Wong, T. Y., Smith, W., Klein, R., & Leeder, S. R. (2004). Retinal microvascular signs and risk of stroke and stroke mortality. Neurology, 62(7), 1062–1068.

[11] Poplin, R., Varadarajan, A. V., Blumer, K., et al. (2018). Prediction of cardiovascular risk factors from retinal fundus photographs via deep learning. Nature Biomedical Engineering, 2(3), 158–164.

[12] Rim, T. H., Lee, G., Kim, Y., et al. (2020). Prediction of systemic biomarkers from retinal fundus photographs: a multi-task deep-learning approach. The Lancet Digital Health, 2(6), e297–e305.

[13] Squirrell D, Yang S, Xie Li, Ang S, Moghadam M, Vaghefi E, McConnell MV. Blood Pressure Predicted from Artificial Intelligence Analysis of Retinal Images Correlates with Future Cardiovascular Events. JACC: Advances 2024, 3;12: 101410. 10.1016/j.jacadv.2024.101410

[14] Vaghefi, E., Squirrell, D., Yang, S., et al. (2024). Development and validation of a deep-learning model to predict 10-year atherosclerotic cardiovascular disease risk from retinal images using the UK Biobank and EyePACS 10K datasets. Cardiovascular Digital Health Journal, 5, 59–69.

[15] Vaghefi, E., An, S., Corbett, R., & Squirrell, D. (2024). Association of retinal image-based, deep learning cardiac BioAge with telomere length and cardiovascular biomarkers. Optometry and Vision Science, 101(7S), e2158.

[16] Sudlow, C., Gallacher, J., Allen, N., et al. (2015). UK Biobank: An Open Access Resource for Identifying the Causes of a Wide Range of Complex Diseases of Middle and Old Age. PLOS Medicine, 12(3), e1001779.

[17] Jones DW, Ferdinand KC, Taler SJ, et al 2025 AHA/ACC/AANP/AAPA/ABC/ACCP/ACPM/AGS/AMA/ASPC/NMA/PCNA/SGIM Guideline for the Prevention, Detection, Evaluation and Management of High Blood Pressure in Adults: A Report of the American College of Cardiology/American Heart Association Joint Committee on Clinical Practice Guidelines. J Circulation. doi:10.1161/CIR.0000000000001356. https://www.ahajournals.org/doi/abs/10.1161/CIR.0000000000001356

[18] Flammer, J., Konieczka, K., Bruno, R. M., et al. (2013). The eye and the heart. European Heart Journal, 34(17), 1270–1278.

[19] Ding, J., Wai, K. L., McGeechan, K., et al. (2014). Retinal vascular caliber and the risk of stroke: a meta-analysis. Neurology, 82(9), 782–789.

[20] Mitchell, P., Leung, H., Wang, J. J., et al. (2005). Retinal vessel diameter and the risk of hypertension: the Blue Mountains Eye Study. Journal of Hypertension, 23(2), 275–281.

[21] Schiffrin, Ernesto L. How Structure, Mechanics, and Function of the Vasculature Contribute to Blood Pressure Elevation in Hypertension. Canadian Journal of Cardiology, Volume 36, Issue 5, 648–658

[22] Humphrey JD. Mechanisms of Vascular Remodeling in Hypertension. Am J Hypertens. 2021 May 22;34(5):432–441. doi: 10.1093/ajh/hpaa195. PMID: 33245319; PMCID: PMC8140657.

[23] Sharrett, A. R., Hubbard, L. D., Cooper, L. S., et al. (1999). Retinal arteriolar diameters and risk of hypertension: the Atherosclerosis Risk in Communities (ARIC) Study. American Journal of Epidemiology, 150(3), 263–270.

[24] Global Cardiovascular Risk Consortium. Global Effect of Cardiovascular Risk Factors on Lifetime Estimates. N Engl J Med. 2025 Jul 10;393(2):125–138. doi: 10.1056/NEJMoa2415879. Epub 2025 Mar 30. PMID: 40162648.

[25] Vaghefi E, An S, Moghadam S, Yang S, Xie L, Durbin M etal. Retinal BioAge Reveals Indicators of Cardiovascular-Kidney-Metabolic Syndrome in US and UK Populations. 2024/7/19. medRxiv. 2024.07. 18.24310670

[26] Tufail, A., Rudisill, C., Egan, C., et al. (2017). Automated diabetic retinopathy image assessment software: diagnostic accuracy and cost-effectiveness compared with human graders. Ophthalmology, 124(3), 343–351.

[27] Bhuiyan, A., Govinda, U., & Papodopoulous, M. (2020). Retinal fundus image analysis for screening and diagnosis of systemic and ocular diseases. Journal of Clinical Medicine, 9(4), 1149.

[28] Marston, H. R., & Freeman, E. E. (2021). Heart age, a novel component of cardiovascular disease risk communication: A qualitative study of the opportunities for clinical implementation. BMJ Open, 11(4), e044439.

[29] Fryar, C. D., Chen, T. C., & Li, X. (2012). Prevalence of uncontrolled risk factors for cardiovascular disease: United States, 1999–2010. NCHS Data Brief, (103), 1–8.

